# Enforced inactivity in the elderly and diabetes risk: initial estimates of the burden of an unintended consequence of COVID-19 lockdown

**DOI:** 10.1101/2020.06.06.20124065

**Authors:** Courtney Kipps, Mark Hamer, Neil Hill, Paula Lorgelly

## Abstract

**Background:** Older adults and those with underlying health conditions were advised to stay at home to help reduce the spread of COVID-19 however little advice on regular physical activity was given to those at risk. We modelled the effects of enforced inactivity on diabetes burden using published evidence.

**Methods:** Using Health Survey for England data, we estimated the prevalence of pre-diabetes and physical activity in adults aged 70 and older. The number of new diabetes cases directly attributed to lockdown were calculated using population attributable risk. Unit cost estimates of the additional burden on primary care and the cost of complications to secondary care were taken from the literature.

**Results:** From 9 million older (≥70yrs) people living in England, 2.1 million could be defined as pre-diabetic (glycated haemoglobin 42<48 mmol/mol). The estimated population attributable fraction (0.281) (assuming relative risk of diabetes from inactivity=3.3, 40% physically active) would give rise to 392,948 new cases of diabetes which we argue are directly attributed to a prolonged period of lockdown. We estimate that the cost of screening and testing these patients in primary care (£35m), their subsequent treatment and management (£229m), and complications (£909m) would equate to an additional £1.17bn to the health care system.

**Conclusions:** Inactivity related to lockdown in previously active older adults may contribute up to £1.17b in additional healthcare costs through a potential increase in diabetes. Clear advice about the importance of physical activity may reduce this potential economic burden during global pandemics.

## Introduction

Governments around the world have imposed a lockdown on their residents in a coordinated effort to control the COVID-19 infection rate. In late March the United Kingdom government passed legislation to impose restrictions on people’s movements and activities. These regulations stipulated that people would not be allowed to leave their home other than for certain specific reasons which included buying food, seeking health care and to exercise once a day. People with chronic underlying health conditions and those aged 70 years or older, known to be at high risk if they should catch coronavirus, were advised to stay at home for at least 12 weeks.

Those shielding at home were instructed to avoid all contact with others, except for essential medical treatment or support. Many older adults voluntarily chose to practice strict isolation to keep themselves safe. These measures, alongside the closure of leisure and community facilities, enforced limitations on physical activity that appeared to be particularly evident in those at greatest risk.(1)

Physical activity is a key factor in the prevention and treatment of a multitude of chronic health conditions. Those who are the most physically active have the lowest rates of chronic disease(2,3) and the lowest mortality,(4) remain more productive for longer(5) and are associated with subsequently lower healthcare costs.(5–7) Conversely, those who are the least physically active have higher rates of chronic disease which contributes to a greater reliance on healthcare systems and higher healthcare costs to society. The health benefits of regular activity can be lost surprisingly quickly. For example, withdrawal from exercise for 1-2 weeks can result in physiological effects such as changes to glucose homeostasis.(6,7) Thus, it is plausible that 12 weeks of inactivity may exacerbate existing conditions in those with risk factors.(8,9)

The indirect health effects of lockdown have been widely discussed in both popular and scientific press, often concentrating on reduced attendance in emergency departments and general practice.(10) There has been little discussion on the adverse effects of isolation itself on physical health.(11)

Type 2 diabetes is a major risk factor for cardiovascular disease, renal failure and stroke. In the United Kingdom, treatment for type 2 diabetes and its complications costs the NHS around £8.8 billion per year.(12) The prevalence of diabetes in England has risen from 2.4% to 6.9% between 1994 and 2016.(13) There is strong evidence that physical activity, along with weight loss and a healthy diet, is integral to reducing the risk of developing diabetes.(14–17)

This paper seeks to estimate the effect of inactivity during lockdown on diabetes risk in the elderly, and the subsequent healthcare costs of managing an increased incidence of diabetes and its associated complications.

## Method

Diabetes is often preceded by pre-diabetes, more recently referred to as non-diabetic hyperglycaemia (NDH), and/or impaired glucose tolerance. Pre-diabetes refers to raised blood glucose levels below the reference range for diabetes and includes impaired fasting glucose (defined as glucose levels between 6.0-6.9mmol/L. by the World Health Organisation(18) and 5.5-6.9mmol/L by NICE(19) and the UK NHS Diabetes Prevention Programme(20) and/or glycated haemoglobin [HbA1c] values between 42-47mmol/mol (6.0-6.4%)).

To estimate the additional incident cases of diabetes in the elderly due to lockdown, we chose to focus on those elderly with pre-diabetes, thereby indicating they are at high risk of developing type 2 diabetes.(21) Using an identical approach to that previously employed by Public Health England(22) but with more recent Health Survey for England (HSE) data, 17.3% of adults aged 70 years or older in England can be classified as having NDH, that is have pre-diabetes. Some of these individuals would have progressed to diabetes regardless of COVID-19, thus we assumed a background conversion rate of 10%(23). Our “at risk” group therefore represents the 90% of elderly with pre-diabetes who are unlikely to have progressed to diabetes within the year.

### Effect estimates for inactivity and risk of diabetes

To determine the effect of inactivity imposed by lockdown on the active elderly, we sought to estimate the proportion of the elderly population with pre-diabetes who are physically active and estimate their relative risk of diabetes.

Using data from the HSE (pooled for 2013-2017 to ensure a large enough sample) we estimated the proportion of active elderly with HbA1c of 42-47mmol/mol (6.0-6.4%), who reported at least 150 minutes of moderate to vigorous physical activity (MVPA) in the previous week (note that this definition aligns with the national guidelines(24)). From this we assumed that the active elderly at risk of diabetes - the prevalence of exposed cases - was 40.3% (this estimate is similar to the estimates for the overall elderly population(25)).

To estimate the effect of inactivity on diabetes risk we used data from a large diabetes prevention programme that compared metformin to a lifestyle intervention in individuals at risk of diabetes.(16) As the trial looked at the effect of increasing physical activity it was necessary to estimate the inverse effect of reducing physical activity by calculating the relative risk of inactivity on conversion rates to diabetes. Knowler *et al*. reported 43 new diabetes cases over 1 year in the exercise group (n=1079), and 141 new diabetes cases in sedentary control (n=1082).(16) Assuming the sedentary controls (i.e. the metformin group) are treated as the ‘exposed’ and the lifestyle group are treated as the ‘control’ we estimated a relative risk (RR) of 3.3 (95% CI: 2.3, 4.5).^1^ A more conservative approach using data from multiple trials gives a near identical RR (3.3, 95% CI: 2.4,4.4).(14) This relative risk allowed us to estimate the population attributable fraction (PAF) given the proportion of elderly who are active (the prevalence of exposed cases).(26) We estimated the PAF to be 0.281, e.g. 28.1% of elderly with pre-diabetes would be likely to develop type 2 diabetes over the next 12 months due to the effect of transitioning into inactivity during lockdown.^2^

### Cost estimation

The approach above allowed us to estimate the number of new cases of diabetes in the elderly due to lockdown. In order to understand the cost of these we used estimates from Hex *et al*.(27) Hex and colleagues used a top-down approach to estimate the total current and future burden of both type 1 and type 2 diabetes. Using their prevalence estimates from 2010/11 we estimated the unit cost of the additional burden on primary care, e.g. the cost of screening and testing and subsequent treatment and management, and the cost of complications to secondary and tertiary care providers. We inflated these costs to 2018/19 prices.(28) As a minimum we would expect newly diagnosed elderly individuals with diabetes to be tested and screened and then managed in primary care. Some proportion of these individuals will develop complications, as modelled in the Hex *et al*. estimates.(27) Combining these costs with the new cases of diabetes provides an estimate of the additional cost of diabetes due to inactivity during lockdown.

### Sensitivity analysis

We tested the sensitivity of our estimates by including the confidence interval bounds on the RR in our analysis. Additionally we tested our assumption of inactivity during lockdown by modelling the effect of more conservative estimates: 25% of the elderly remain active, 50% remain active and 75% remain active.

Table 1 presents the various parameters used in the analysis.

**Table 1:** Parameter values and sources.

| Parameter | Value | Source |
| --- | --- | --- |
| Proportion of individuals 70+ with HbA1c values between 6.0–6.4% (i.e. pre-diabetes) | 17.3% | Authors' own analysis of HSE data 2013-2017 |
| Proportion of individuals who will progress to diabetes irrespective of pandemic | 10% | Tábak *et al.* (23) |
| Proportion of individuals 70+ who are physically active with HbA1c values between 6.0–6.4% | 40.1% | Authors' own analysis of HSE data 2013-2017 |
| Relative risk of inactivity on conversion rates to diabetes | 3.3 | Knowler *et al.*(16) and authors' own calculations |
| PAF of those who were active prior to lockdown who go on to develop type 2 diabetes due to inactivity | 28.1% | Authors' own calculations |
| Total screening and testing cost per patient | £88.95 | Hex *et al.*(27) (inflated) |
| Total management and treatment cost per patient | £582.38 | Hex *et al.*(27) (inflated) |
| Total complication cost per patient | £2312.45 | Hex *et al.*(27) (inflated) |

## Results

The Office of National Statistics estimated that there were 9,006,762 individuals aged 70 or over in the UK in mid-2019.(29) We estimated that 1.554 million over 70 years olds in the UK have HbA1c values that make them at high risk of diabetes, e.g. they have pre-diabetes. In our models we made the assumption that Government advice to stay at home, and other lockdown measures has resulted in the active elderly with pre-diabetes becoming inactive, and as a result of this inactivity up to 392,948 new cases of diabetes in the 70+ age group may be expected.

Screening and testing these patients in primary care is estimated to cost £35m, and their subsequent treatment and management in primary care including prescribing antidiabetic drugs will cost £229m. The largest cost component of these individuals progressing to diabetes is the cost of complications which is estimated to be up to £909m for this newly diagnosed cohort of patients with diabetes. The total cost of inactivity on the NHS is estimated to be £1.17 billion; notably this does not include indirect costs, social care costs or lost productivity due to mortality.

The results of various sensitivity analyses are presented in Table 2. Our most conservative estimate suggests that despite lockdown and government advice 75% of the elderly remain active and only 98,237 transition to diabetes at a cost of £586 million. At the other extreme if lockdown has constrained activity and the relative risk of diabetes conversion due to inactivity is higher, then 438,507 elderly individuals will develop diabetes at a cost of £1.3 billion.

**Table 2:**
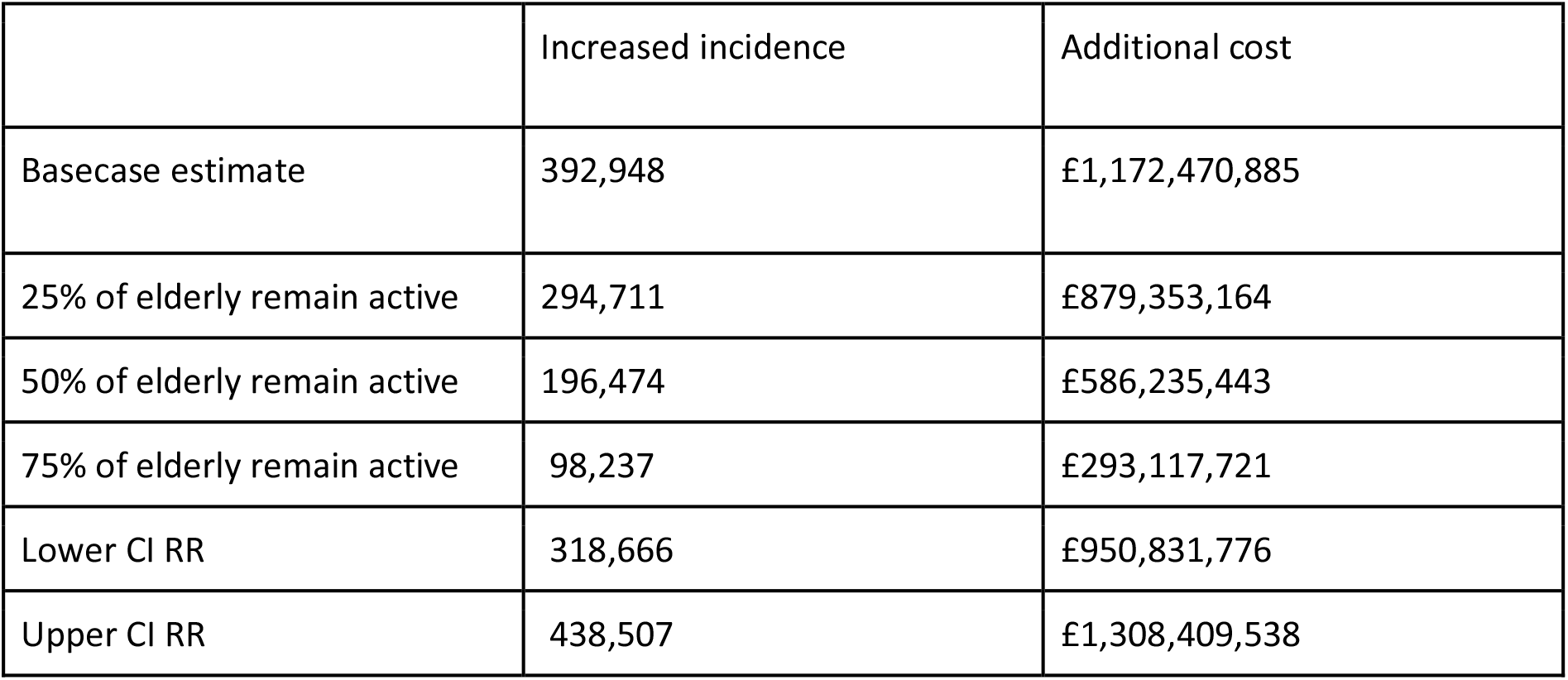
Sensitivity analysis.

## Discussion

We estimate that the excess healthcare costs related to new diagnoses of diabetes in previously active adults over 70 years of age in England who have become inactive due to the COVID-19 lockdown may exceed £1.17 billion.

The costs to the National Health Service of treating patients with COVID-19 are predicted to run into the billions of Pounds. The indirect healthcare costs of COVID-19 pandemic are likely to be substantial but are as yet unknown. Numerous unintended healthcare consequences of COVID-19 have begun to emerge, including excess mortality due to early discharge,(30) widespread delays to routine operations(31), reduced attendance to Emergency Departments(32), and delays in diagnosing serious illness(33) either because people are reluctant to seek medical attention or because normal services (including screening) have been suspended. Each will carry a significant indirect cost burden to be met by the government and it is likely that the breadth of the impact of COVID-19 across the economy will mean cost-savings will need to be made across the NHS. A challenge will be to determine where these can be made.

The annual cost of treating type 2 diabetes account for nearly nine per cent of the NHS budget.(12) Every year around 200 000 people develop diabetes in the UK.(12) The findings from this current study suggest a possibility that at least 98,000 additional new cases of diabetes may occur as a result of the lockdown (applying conservative estimates) and the potential for up to 390,000 new cases. This latter estimate represents a doubling of the annual incidence of diabetes and an additional £1.17 billion in healthcare costs as an indirect effect of COVID-19.

Diabetes is a preventable condition, mainly through behavioural interventions such as exercise and a healthy diet.(16,17,34) During lockdown, exercise has been widely discussed in the media in the context of allowing people to leave their homes, but rarely in the context of prevention of chronic disease. Changing the emphasis towards encouraging exercise for the maintenance of good health and the prevention of chronic diseases may slow this conversion of pre-diabetes to diabetes. Additionally, there is considerable evidence that shows that physical activity is a cost effective means of preventing and managing a range of long term conditions,(2) thus physical activity, and the promotion of physical activity, can play a role in generating much needed efficiency gains in a post-COVID healthcare system. It is important to highlight that exercising at home is compatible with government lockdown regulations.

It is probable that inactivity due to lockdown will have similar effects on other chronic diseases. We used diabetes conversion from pre-diabetes as a model to demonstrate the potential health economic effects because it is a common condition with a well described precursor syndrome(23), clear evidence of risk reduction with physical activity,(34) and widely used cost analysis.(27) These figures may well be replicated across other exercise-dependent chronic conditions including cardiovascular disease and obesity, as well as certain cancers and mental health conditions.

Governments can learn from this pandemic in their preparations for the possibility of similar situations in the future.(35) Simple measures and clear advice about the importance of maintaining physical activity may reduce a potential and unintended health economic burden during future global pandemics in which older vulnerable adults are required to stay at home.

### Assumptions and limitations

This study makes several assumptions. The first assumption is that most adults over 70 years have reduced or even stopped exercise during the lockdown. Whilst some have found ways to exercise safely outdoors, emerging data has found that many either have limited access to open spaces or are concerned about venturing outside of the home where they have a higher chance of encountering the coronavirus.(1)

The second assumption is that inactivity as a result of lockdown will last for at least 12 weeks, if not longer, and that this is sufficient time to observe adverse physiological effects of inactivity on glycaemic control. Although there are currently cautious moves to allow people more opportunities to leave home,(36) it is likely that older adults will be advised to remain at home for longer than those of working age. When restrictions are eventually lifted, it is likely that many of those over 70 will remain cautious about going outside to exercise(36) and thus the effects of lockdown on physical activity in this group may persist beyond the formal end of lockdown regulations.

Alongside physical activity,(34) a healthy diet is key to reducing the risk of developing diabetes, and most diabetes prevention trials include dietary intervention in combination with structured exercise and weight loss.(14–17) Our risk estimates of inactivity on diabetes are drawn from such trials, thus it is difficult to tease apart the effects of diet and weight loss from exercise. This study assumes that calorie consumption through diet and alcohol during lockdown is unchanged compared to pre-lockdown. We have not made any assumptions about weight gain. The true dietary effects of lockdown are multifactorial, and it will be some time before the effects of closing restaurants and cafes and limiting access to supermarkets on developing diabetes will become clear.

We have assumed that the risk of developing diabetes is evenly spread across the population of adults over 70 years of age, and that their subsequent risk of complications is similarly evenly spread. We acknowledge that those who were previously active and who develop diabetes related to inactivity are less likely to suffer early complications than those who were previously inactive, however the datasets which describe these figures are not available.

Despite these limitations our approach is sufficiently robust to enable a useful estimate of the health and cost implications of inactivity during lockdown.

## Conclusion

The unintended and indirect healthcare costs of coronavirus stay-at-home lockdown regulations may be significant. Inactivity in previously active older adults may contribute up to £1.17 billion in additional healthcare costs through a potential increase in the incidence of diabetes. Simple measures and clear advice about the importance of physical activity in vulnerable adults may reduce this potential economic burden during global pandemics in which populations are required to stay at home. Furthermore, this advice should promote that exercise at home is compatible with government lockdown regulations both now and for the foreseeable future.

## Data Availability

no data created

## Footnotes

1 Relative Risk = 141 (43+1036) / 43 (141+941) = 3.3

2 PAF = pc(1 − 1/RR) = 0.403*(1 – (1/3.3)) = 0.281. This is for 12 months as estimated from the relative risk estimate.

